# MOdulation-Guided ENcoding (MOGEN) Scheme for Vessel-Encoded Arterial Spin Labeling

**DOI:** 10.1101/2025.07.23.25332076

**Authors:** Hongwei Li, Thomas W. Okell, Joeseph G. Woods, Yang Ji, Yuriko Suzuki, Tiansheng Qian, Yujun Liao, Jian Wang, Ziqi Sun, Ying-Hua Chu, Yi-Cheng Hsu, He Wang, Zhensen Chen

## Abstract

Vessel-encoded arterial spin labeling (VEASL) enables simultaneous, non-contrast imaging of multiple vascular territories that is useful for differential diagnosis and treatment monitoring of cerebrovascular diseases. However, the existing encoding methods are either signal-to-noise ratio (SNR) inefficient or requiring the spatial modulation to approximate a cosine function. To address these limitations, we developed a MOdulation-Guided ENcoding (MOGEN) scheme that directly exploits the spatial modulation pattern to obtain SNR-efficient encoding matrix. Simulation studies demonstrated that MOGEN achieves significantly higher theoretical SNR efficiency than previous methods across both four-and six-artery configurations. In healthy volunteers, MOGEN improved in vivo SNR by approximately 15% and provided more robust vessel decoding, particularly when the spatial modulation deviated from the cosine profile. In patients with Moyamoya disease, MOGEN enabled reliable visualization of collateral pathways even when scan time was reduced to ∼5 minutes for six arteries. Furthermore, by incorporating vessel size information and ensuring sharp label/control transitions, MOGEN enhanced single-artery selectivity in vessel-encoded angiography. We also demonstrated that a straightforward approach of off-resonance correction for VEASL at ultra-high field was feasible by using MOGEN.

## 1 INTRODUCTION

It is essential to distinguish the vascular territories of the different brain feeding arteries for cerebrovascular diseases^1^, such as the assessment of collateral circulation^2,3^. The vessel-selective information not only facilitates the decision-making in the treatment of cerebrovascular diseases, e.g. arteriovenous malformation^4^ and Moyamoya disease^5^, but also aids in monitoring disease progression^6^. Conventional X-ray digital subtraction angiography (DSA) is considered the gold standard for identifying the arterial source of blood supply in clinical diagnosis, but has obvious limitations including invasiveness and ionizing radiation exposure^7^.

Vessel-selective arterial spin labeling enables non-invasive acquisition of territorial perfusion maps and angiograms in the brain^8^, achieved through two main techniques^9^: super-selective arterial spin labeling (SSASL)^10^ and vessel-encoded arterial spin labeling (VEASL)^11^. SSASL allows for the labeling of a single vessel in each scan without the need for vessel-decoding and offers more flexibility in choosing labeling regions. However, its signal-to-noise ratio (SNR) efficiency is significantly lower than that of VEASL when multiple arteries are of interest^12^, and the scan time increases proportionally with the number of feeding arteries, potentially reaching 10–15 minutes for perfusion territory imaging of six arteries^6^. VEASL instead generates a periodic variation in labeling efficiency within the labeling plane, and a series of images are acquired using different encoding combinations to calculate the amount of blood arising from each feeding artery within each voxel. VEASL has been shown to achieve a SNR comparable to that of standard pseudo-continuous arterial spin labeling (PCASL)^13^. Therefore, VEASL enables the labeling of multiple arteries, such as the nine vessels above the circle of Willis, in approximately 5 minutes without significant compromise of image quality^14^.

Several encoding strategies have been proposed for VEASL. The standard approach labels the four main brain feeding arteries in the neck, i.e. bilateral internal carotid arteries (ICAs) and vertebral arteries (VAs), by using empirical SNR-efficient encodings^13^, including left-right (LR), anterior-posterior (AP), and diagonal encoding pairs. However, this approach is limited to the four-vessel setting, with the vessels approximately located at the corners of a rectangle, which is not always available. Planning-free VEASL usually also adopts several fixed encodings^15^, with assumed locations and relative distances for the vessels, and uses k-means clustering for decoding. However, this approach is primarily designed for three or four vessels in the neck, and may offer insufficient vessel selectivity contrast^16^, resulting in a limited ability to detect mixed perfusion, even with the Bayesian inference framework^16^. Random encoding is an alternative approach which utilizes many pre-defined vessel encoding cycles without the need for precise planning^17^, but the SNR efficiency is significantly lower than that of the standard approach for four arteries^18^. The high number of encoding cycles is also unsuitable for vessel-encoded angiography due to the long acquisition time for each encoding cycle.

The optimized encoding scheme (OES) is currently the landmark for achieving optimal SNR in VEASL for an arbitrary number of vessels and vessel locations^18^. For each pre-defined encoding pattern, OES produces the optimal encoding parameters (i.e. α, λ, ε, as defined below) by identifying the maximum point on the Fourier transform of the ideal encoding pattern in the labeling plane. However, OES essentially assumes that the spatial modulation of VEASL labeling approximates a cosine function. Such an assumption makes OES valid only under certain PCASL labeling parameter settings of the bipolar approach with smoother cosine function-like transitions between label and control conditions, even though this is not always the case^19^. In addition, the OES does not optimize the combination of the encoding patterns. This issue was addressed by the improved OES (IOES) by minimizing the condition number of the actual encoding matrix and the sensitivity to motion^14^. Another shortcoming of both the OES and the IOES is that during the optimization, each vessel is represented as a point on the labeling plane and actual size of the vessel is ignored.

As such, in this study, we propose a rapid modulation-guided encoding (MOGEN) scheme for VEASL to effectively consider the spatial modulation under arbitrary PCASL parameters settings and further optimize the SNR. To demonstrate the capability of MOGEN, we primarily focus on the simultaneous encoding of intra-and extracranial arteries, targeting six main feeding arteries in the neck, and validate its effectiveness in Moyamoya disease. Additionally, we evaluate the feasibility of incorporating the vessel size information into MOGEN to achieve highly-selective angiograms by combing with the unipolar approach. Furthermore, we proposed the framework to perform off-resonance correction for MOGEN-based VEASL and showcase its feasibility with 7T experiments.

## 2 THEORY

### 2.1 Encoding Schemes

The VEASL signal of each voxel across encoding cycles, **y**, can be described as,

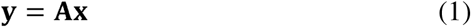

where **A**^+^ is the vessel-encoding matrix, describing the modulation of the signal from each artery and static tissue across the encoding cycles, and **x** is a vector of contributions to the total signal from each arterial component plus static tissue. The goal of VEASL is to recover **x** from **y** as efficiently as possible by designing an encoding matrix **A** that has a low condition number and high SNR efficiency, which can be defined as follows^11^:

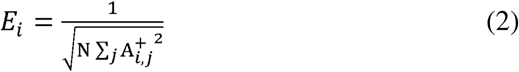

where **A^+^** is the pseudoinverse of **A**, N is the number of columns in **A^+^**. Ideal encoding matrices are those that achieve the SNR efficiency of one for all vessels^18^, which necessitates an equal number of perfect label (-1) and control (+1) conditions for each artery. Such encoding matrices would be composed of columns derived from a Hadamard matrix.

### 2.2 The MOGEN Method

Similar to OES and IOES, the MOGEN method is designed to identify the optimal encoding matrix as well as the VEASL imaging parameters, given the number and locations of the target arteries. However, MOGEN abandons the assumption of a cosine spatial modulation and can yield the optimal encodings under arbitrary shape of spatial modulation, which in turn improves the flexibility of the PCASL parameters for VEASL.

Given a set of PCASL parameters, including RF flip angle (FA), maximum gradient amplitude (G_max_), mean gradient amplitude (G_mean_), RF duration (RFdur), and the interval between two RF pulses (RFsep), and the blood velocity, the nominal spatial modulation of the spins in one full cycle (i.e. the phase accrued between consecutive PCASL pulses spanning from-π to π), with control defined at a phase of 0, and label at phases of π and - π, are calculated by Bloch equation simulations. **Figure 1a** shows the simulated spatial modulations under two sets of PCASL parameters and different blood velocities. In this study, all simulated spatial modulations used in the calculation of MOGEN were obtained with a velocity of 30 cm/s^20^.

**Figure 1.**
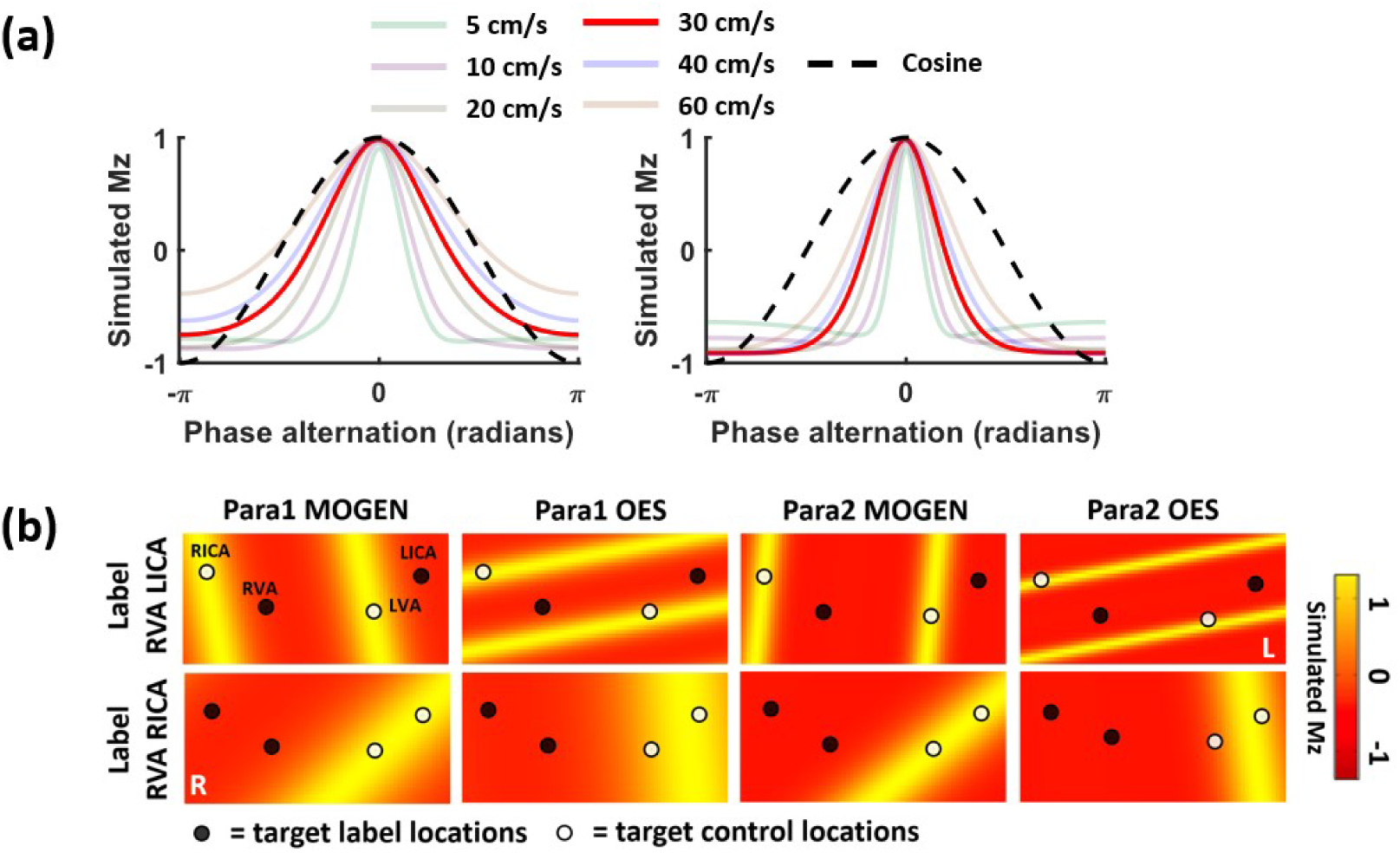
(a) Simulated spatial modulation of labeling for PCASL parameter set 1 (Para1, left) and 2 (Para2, right). The spatial modulation of Para1 closely approximates a cosine function under a velocity of 30 cm/s, while Para2 shows higher labeling efficiency and is less sensitive to flow velocity; (b) The resulting encoding designs by MOGEN and OES under Para1 and Para2 for an example set of artery locations and target encoding schemes (i.e. “Label” for RVA and LICA, “Control” for LVA and RICA in the first row; “Label” for RVA and RICA, “Control” for LVA and LICA). The LVA deviates from the desired “Control” state being more pronounced in Para2 using OES. Para1: FA of 20°, G_max_ of 6.0 mT/m, G_mean_ of 0.8 mT/m, RFdur of 500 μs, and RFsep of 1000 μs; Para2: FA of 28°, G_max_ of 5.0 mT/m, G_mean_ of 0.36 mT/m, RFdur of 480 μs, and RFsep of 1210 μs.

Figure 2 shows the optimization algorithm framework of MOGEN, exemplified with a 6-artery and 8-cycle setting. In VEASL, the labeling pattern on the labeling plane is determined by 3 parameters: the angle (α), wavelength (λ), and the offset relative to the magnet isocenter (ε). The optimization in MOGEN involves searching the optimal (α, λ, ε) for each encoding row in the matrix **A**, and the optimal combinations of encoding columns, extracted from a Hadamard matrix, to form **A**. The implementation details of MOGEN are as follows:

**Figure 2.**
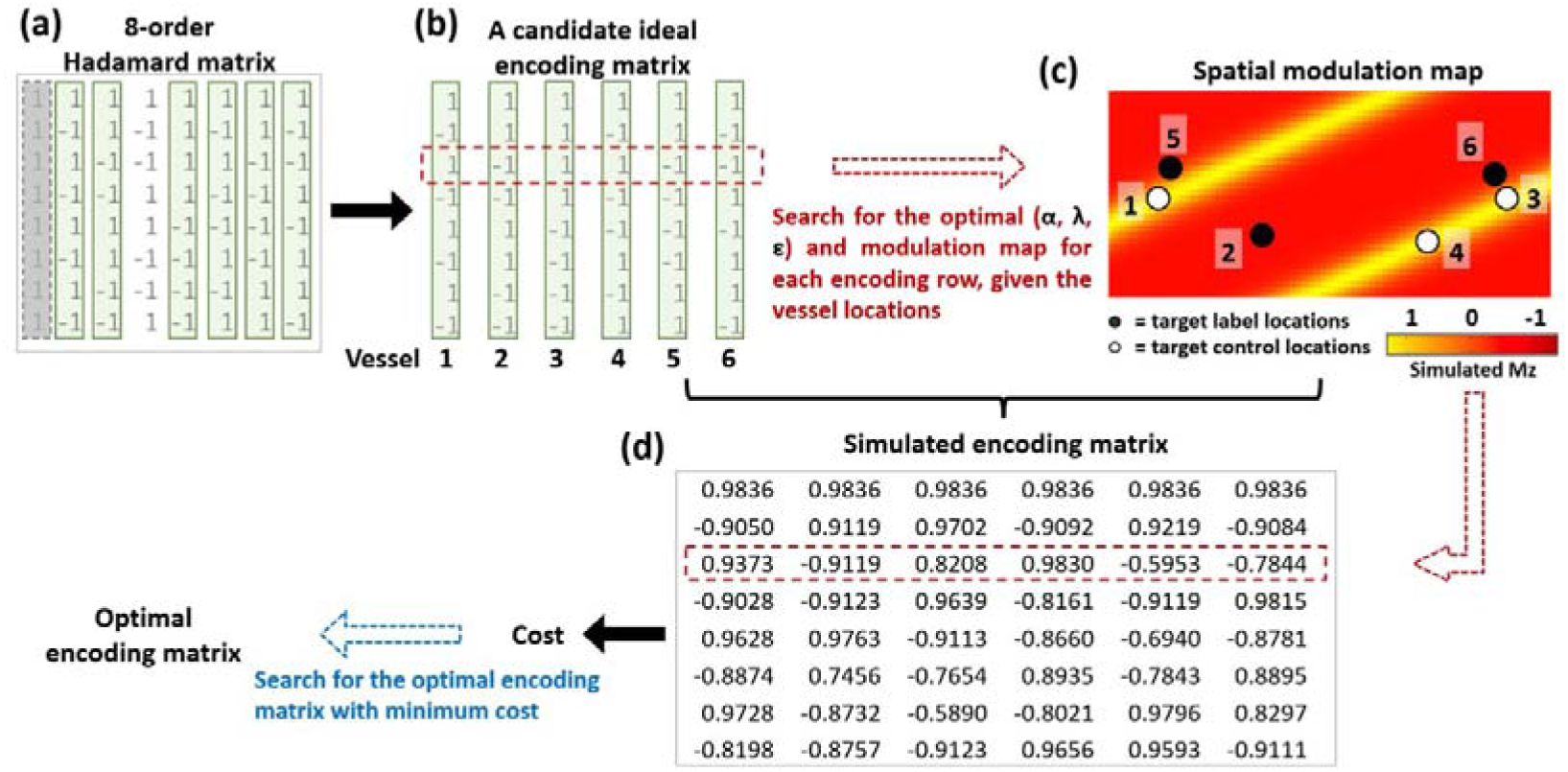
The optimization framework of MOGEN for identifying the optimal encoding matrix and the labeling parameters (α, λ, ε), exemplified with a 6-artery and 8-cycle setting. (a) For 6 target vessels, the minimum Hadamard matrix order (size 8) exceeding 6 was selected. (b) All 6-column sub-matrices were extracted from the Hadamard matrix as encoding matrix candidates. (c) Target vessel values from modulation maps were extracted, and dot products with ideal encoding matrix rows were calculated. The map yielding the highest dot product per row defined optimal (α, λ, ε). (d) The optimal (α, λ, ε) from all rows generated a simulated encoding matrix for each matrix candidate, followed by the cost function to select the final encoding matrix.

#### Generation of candidate ideal encoding matrices

Given N target vessels, the Hadamard matrix’s minimum order that is larger than N is determined (Figure 2a). The column containing all ones in the Hadamard matrix (corresponding to static tissue) is removed. Then all sub-matrices containing N columns are extracted from the remaining part of the Hadamard matrix, with each sub-matrix being a candidate of the encoding matrix **A** (Figure 2b).

#### Generation of spatial modulation maps

In MOGEN, the labeling plane is represented as a 1024×1024 matrix and a spatial modulation map on the plane is generated for each (α, λ, ε) based on the nominal spatial modulation under a set of PCASL parameters (Figure 2c). A dictionary of spatial modulation maps was generated for all potential (α, λ, ε). In this study, when generating the dictionary, we let α span from 0° to 359° in 1° increments, and λ span from the shortest intervascular distance between target vessels to a maximum of 150 mm in 2 mm increments, and ε span from-75 mm to 74 mm in 1 mm increments.

#### Determination of the optimal (α, λ, ε) for each encoding row

On each modulation map, the values at locations of the target vessels are extracted. Then the dot products of these values with each row of an ideal encoding matrix are calculated. For each encoding row, the modulation map yielding the highest dot product is determined, with the corresponding (α, λ, ε) deemed optimal (Figure 2c).

#### Selection of the optimal ideal encoding matrix

For each candidate ideal encoding matrix, once the optimal (α, λ, ε) parameters are identified across all encoding rows, a simulated encoding matrix is constructed by incorporating target vessel values from the modulation maps (Figure 2d). Then, similar to IOES^14^, the optimal encoding matrix is identified by minimizing the cost in **Equation 3**:

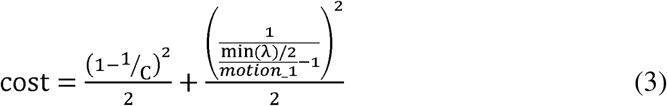

where C is the condition number of the simulated encoding matrix, min(λ) is the minimum wavelength and *motion_*1 is a constant (4 mm in this study) that controls motion sensitivity.

#### Representation of vessels

In MOGEN, each vessel is represented as either a single voxel or multiple voxels on the spatial modulation maps. The later allows the consideration of the vessels’ size during the optimization. In the following part of this paper, the MOGEN with a single-voxel and multi-voxel vessel representation are referred to as MOGEN and MOGEN_V2, respectively.

#### Other features of MOGEN

Motion occurring along the offset dimension (i.e. the direction of the transverse gradient) will lead to shift of the spatial modulation and potential wrong labeling, especially when the spatial modulation has a sharp control-to-label transition or a narrow control (or label) band. To consider this effect, a weighted averaging was applied to the dictionary of spatial modulation maps along the offset (ε) dimension using the weighting function *H* as defined in **Equation 4**, with *motion_*2 being an adjustable parameter (4 mm in this study).

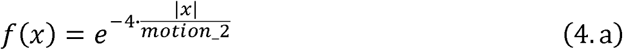

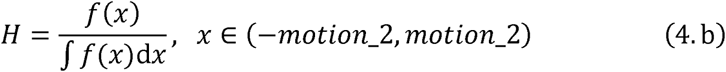

To accelerate MOGEN calculation, only the values at around the vessel locations are calculated when generating the spatial modulation maps. In addition, during the ergodic search, the optimal (α, λ, ε) and the corresponding simulated encoding values for a specific encoding row are calculated only once and reused when the same row appears in other encoding matrix.

### 2.3 The Off-Resonance Correction

When B_0_ inhomogeneities are present, additional phase accumulation during the RF intervals of the PCASL pulse train leads to phase offsets, which will undermine the labeling efficiency and cause the spatial modulation of VEASL to deviate from the simulation. The unipolar approach with consistent gradient polarity during labeling, could be applied to counteract the effects of off-resonance^17^.

The off-resonance correction for MOGEN-based VEASL is straightforward. Specifically, the local phase offsets Δ< at the vessel position of interest is subtracted out from the original phase < when generating the spatial modulation maps. The phase offsets can be measured through a single voxel corresponding to the vessel center on the phase difference map from a single-slice dual-echo field map at the labeling plane^21^.

## 3 METHODS

### 3.1 Overview

The SNR efficiency of the MOGEN method was compared against OES/IOES through simulations and healthy volunteer experiments under two scenarios. The first scenario included four arteries, bilateral ICAs and VAs using eight Hadamard cycles, referred as 4Ves8Cycles^22^. The second scenario included six arteries, with the two external carotid arteries (ECAs) included as well, and employed 8 (minimum number of encoding cycles) or 12 cycles plus an additional non-selective label for calculating relative labeling efficiency or thresholding the rough gray matter region via the non-selective perfusion signal^12^, referred to as 6Ves9Cycles or 6Ves13Cycles. The MOGEN method was also tested on five Moyamoya patients to demonstrate its clinical potential for the simultaneous encoding of six arteries.

The capability of MOGEN in considering the vessel size was tested by performing in vivo VEASL angiogram scans using MOGEN_V2, MOGEN and OES, with the unipolar VEASL approach adopted. The vessel selectivity across the three scans was then compared.

The effectiveness of MOGEN-based off-resonance correction was evaluated on a phantom at 3T, and in-vivo scans at 7T. The phantom experiments aimed at visualizing the encodings within the labeling plane before and after the off-resonance correction. The in-vivo scans were performed to validate the feasibility of implementing VEASL with MOGEN at ultra-high field, the results of which were also compared with those of the recently proposed dynamic shimming method^23^.

All 3T scans in this study took place on a Siemens 3T Prisma (Siemens Healthineers, Erlangen, Germany) with a 32-channel head coil, while the 7T data were acquired on a MAGNETOM Terra 7T Scanner (Siemens Healthineers, Erlangen, Germany) equipped with an 8-transmit/32-receive head coil. The scans were performed under a technical development protocol agreed by local ethics and institutional committees. Details of the simulations and experiments are described below.

### 3.2 Theoretical SNR Efficiency Comparison

Simulations were performed in MATLAB (MathWorks, Natick, Massachusetts, USA) to compare the theoretical SNR efficiency of the MOGEN and the OES/IOES method. Two sets of PCASL parameters with the bipolar approach were used in the simulations. Para1 followed the default parameters utilized in the OES studies with spatial modulation resembling a cosine function (Figure 1a), which were set as follows^18^: FA of 20°, G_max_ of 6.0 mT/m, G_mean_ of 0.8 mT/m, RFdur of 500 μs, and RFsep of 1000 μs; Para2 was similar to parameters from a previous study, achieving higher labeling efficiency and reduced sensitivity to flow velocity (Figure 1a)^24^: FA of 28°, G_max_ of 5.0 mT/m, G_mean_ of 0.36 mT/m, RFdur of 480 μs, and RFsep of 1210 μs. An arrangement of four or six vessels, representing brain-feeding arteries above the carotid bifurcation in the neck, was used to initialize the coordinates of the simulated vessels.

The first simulation performed was to compare the SNR efficiency of MOGEN and OES/IOES under various configurations of the vessel positions. To imitate the potential variation among individuals, a total of 100 different vessel positions were generated by perturbing the initial coordinates. For the four-vessel arrangement, the perturbations for each vessel in the LR and AP directions were independently selected from a uniform distribution and scaled by half of the shortest distance in the LR and AP directions. The MOGEN and OES were used to generate the 4Ves8Cycles VEASL encodings for each vessel position configuration. For the six-vessel arrangement, the locations of the ICAs were kept fixed, while the positions of the ECAs and VAs were randomized in the same manner as that for the four-vessel arrangement. The MOGEN and IOES were used to generate the 6Ves9Cycles and 6Ves13Cycles VEASL encoding for each vessel position configuration.

The longitudinal magnetization at each vessel location for each encoding cycle in both OES/IOES and MOGEN was directly obtained from the simulated spatial modulation. The SNR efficiency was calculated and averaged across all vessels, and then compared between MOGEN and OES/IOES using paired t-tests.

The second simulation assessed the robustness of MOGEN to head motion compared to OES/IOES. To achieve this, MOGEN and OES/IOES were first applied to calculate the encodings for the initial vessel positions that were the same as the first simulation. Subsequently, the whole labeling plane was shifted in random directions and distances, sampled from a normal distribution with a mean of zero and standard deviations ranging from 0 to 5 mm. The shifting was repeated for 100 times. The longitudinal magnetization of each vessel as well as the SNR efficiency was subsequently calculated and averaged across all vessels and all 100 repetitions.

The calculation time for MOGEN was short: < 1 s for 4Ves8Cycles, < 7 s for 6Ves9Cycles and < 13 s for 6Ves13Cycles. All computations were performed on a system with an 8-core CPU and 128GB RAM.

### 3.3 Phantom Scans

A silicon oil phantom was scanned at 3T to assess the feasibility of off-resonance correction for the MOGEN method. Single-slice VEASL images, overlapping with the labeling plane, were acquired using a spoiled-gradient echo (SPGR) readout. The other imaging parameters were labeling duration = 700 ms, post-labeling delay = 2 ms, voxel size = 1.5×1.5×3 mm³. In the resulting VEASL images, the region with a labeling condition is expected to be manifested as a dark band due to the saturation effect induced by the labeling pulses. Similar to the in-vivo scan as explained below, a fast 3D time-of-flight (TOF) scan was performed, and then coordinates of 4 points (i.e. fake vessels) on the image were extracted for MOGEN calculation. Para1 settings with unipolar approach was used for the VEASL sequence in this experiment.

The VEASL sequence was scanned under three scenarios: (1) third-order standard shimming was used; (2) the X linear shim term was manually reduced by 85 μT/m, introducing phase variations in X direction; (3) the X linear shim was modified as in (2) and off-resonance correction was performed during MOGEN calculation. A single-slice dual-echo field map was used to obtain phase offsets in less than half a minute, with the time interval between the two echoes made equal to the RF separation in Para1, thereby eliminating the need for phase unwrapping and additional calculations (TR = 100 ms, TE1 = 5.07 ms, TE2 = 6.07 ms, voxel size = 1.5×1.5×3 mm³).

### 3.4 In Vivo Healthy Volunteer Study

A total of four in vivo experiments were conducted, with three at 3T and one at 7T.

**Experiment 1**: To assess whether the MOGEN method yields improved SNR, MOGEN was compared with OES/IOES in seven healthy subjects (all female, 28.5±11.8 years) under two scenarios, 4Ves8Cycles and 6Ves13Cycles, utilizing Para1 and Para2 for each condition, resulting in four distinct combinations. The VEASL perfusion scans were performed with the following parameters to generate vascular territory images (VTI): a single-shot echo planar imaging (EPI) readout, labeling duration = 1800 ms, post-labeling delay = 1800 ms, voxel size = 3.4×3.4×5 mm³, pre-saturation plus two global inversion pulses, 10 averages and 6 min 14 s for 4Ves8Cycles, 8 averages and 8 min 5 s for 6Ves13Cycles. An additional dynamic thick-slab 2D projection VEASL angiography scan was performed with bSSFP readout^22^, Para2 and 6Ves13Cycles (labeling duration = 800 ms, post-labeling delay = 2 ms, voxel size = 1.1×1.1×60 mm³, pre-saturation only, total scan time 2 min) to visualize the SNR improvement achieved by the MOGEN method.

**Experiment 2**: Three healthy subjects (2 female, 36.7±15.5 years) were scanned under 6Ves9Cycles and 6Ves13Cycles with Para2, to evaluate the impact of encoding number on the vessel-decoding performance of MOGEN and IOES in the more challenging 6 vessel scenario. Both perfusion and 2D angiography data were acquired, using the same imaging parameters as Experiment 1. The total time for the perfusion scan with 6Ves9Cycles and 6Ves13Cycles were 7 min and 8 min 5 s, respectively. The total time for the 2D angiography scan were 1 min 23s and 2 min, respectively.

**Experiment 3**: MOGEN_V2, MOGEN, and OES were compared to evaluate their single-artery selective capabilities. As for 3D angiography data, since each encoding took more time to acquire, a lower number of encoding cycles would be preferred in clinical applications. This could be achieved by simply targeting a single vessel and reducing encoding cycles along with a less complex vessel-decoding process^19^. Three subjects (3 female, 26.3±0.6 years) were scanned using 3D SPGR dynamic vessel-encoded MRA (unipolar approach, labeling duration = 1000 ms, post-labeling delay = 2 ms, constant flip angle = 10°, temporal resolution = 186.31 ms, TE = 3.56 ms, TR = 6 ms, voxel size = 1.2×1.2×1.2 mm³, pre-saturation only, total scan time 16 min 40 s) with an optimized setting of PCASL parameters^19^, referred to as Para3, designed for a unipolar approach to achieve sharp label/control transitions in spatial modulation, thereby avoiding partial labeling in vessel-selective angiograms^19^. The Para3 were as follows: FA = 20°, G_max_ = 6.0 mT/m, G_mean_ = 0.4 mT/m, RFdur = 500 μs, and RFsep = 1000 μs. Four main feeding arteries were targeted, but the two VAs were treated as a single entity representing the posterior circulation blood supply. Consequently, the encoding scheme comprised five cycles: non-selective control and label, labeling of the RICA, labeling of the LICA, and labeling of both VAs. The control image was subtracted from each vessel-selective encoding image, followed by maximum intensity projection (MIP) along the slice direction, allowing direct visualization of the angiogram quality.

**Experiment 4**: VEASL perfusion data were acquired at 7T to compare dynamic shimming and MOGEN-based off-resonance correction, utilizing a unipolar approach with Para1 in seven subjects (3 female, 24.7±4.4 years). A single-slice field map (TR = 30 ms, TE1 = 4.26 ms, TE2 = 5.32 ms, voxel size = 0.7×0.7×2 mm³, scan time 20 s) was acquired for both methods to correct for B_0_ inhomogeneities within the labeling plane. All VEASL scan protocols were consistent with those in Experiment 1, except for the implementation of variable-rate selective excitation (VERSE) during the labeling to reduce specific absorption rate (SAR) while maintaining SNR^25^. The encoding scheme was the same as that in Experiment 3, i.e. five encoding cycles, and the number of averages was 10, resulting in a total scan time 3 min 51 s.

To ensure a fair comparison, the scan order was randomized in each experiment. A fast 3D TOF scan (1 min 36 s) was always performed at the start for labeling plane selection and vascular coordinates recording. In Experiments 1 and 2, the labeling plane was positioned slightly above the carotid bifurcation. In Experiments 3 and 4, the labeling plane was placed slightly below the confluence of the two VAs, where the planning did not exhibit excessively low B_1_^+^ at 7T and also avoided tissue-air interface where the B_0_ can change rapidly across vessels^26^.

### 3.5 Moyamoya Disease Patient Study

To evaluate the clinical potential of MOGEN in encoding the six main brain feeding arteries, five pre-operative Moyamoya patients (4 female, 47.0±5.9 years) were consecutively recruited and underwent a VEASL perfusion scan. Patient informed consent and institutional ethical and committee approval were obtained in advance for MRI scans. Given the significant variability in blood flow velocity across the six main feeding arteries in Moyamoya patients, Para2 was selected for the 6Ves13Cycles perfusion scan due to its lower sensitivity to velocity variations. To account for the potential slower blood flow in the anterior and collateral circulations, characterized by prolonged arterial transit time, the post-labeling delay was extended to 2000 ms. All other scanning parameters of VEASL remained consistent with those used in healthy volunteers, including 8 averages and a total scan time of 8 min 26 s. The VEASL data were also analyzed using a reduced number of averages (5 averages, scan time = 5 min 18 s) to evaluate whether collateral circulation visualization with VEASL maintained comparable image quality to that obtained with 8 averages, despite a slight reduction in SNR and a scan duration of approximately 5 minutes for six arteries.

### 3.6 Image Analysis

The vessel-decoding analysis was performed using a Bayesian maximum a posteriori (MAP) approach^27^, which was capable of partially addressing partial labeling and poorly conditioned encoding and could also provide information on mixed blood supply, a feature that was quite common in Moyamoya patients^28^. Two vessels per class were used to allow for probabilistic representation of mixed perfusion^12^. Since in healthy volunteers the ECAs are not expected to feed the intracranial region, the accuracy of the VTI images in the healthy subjects was qualitatively examined by checking the decoded territory of ECAs and the presence of mixed perfusion between ECAs and ICAs. For Moyamoya patients, accuracy of the VEASL results could be assessed by comparing with digital subtraction angiography (DSA). However, such a comparison was beyond the scope of this study, and only SNR comparisons (see below) and spatial correlation analyses for each decoded territory between the 5-and 8 average data were conducted. The MAP analysis was not performed in Experiment 3 and 4 as only one vessel was labeled in each encoding cycle.

#### 3.6.1 In Vivo Mean SNR Calculation

The VTI obtained through MAP analysis in Experiment 1 and 2 were used to compare the SNR between MOGEN and OES/IOES. The pipeline detailing the subject inclusion in the two experiments and the number of enrolled participants for SNR comparison was illustrated in **Figure S1**. The signal from the dominant feeding artery in each voxel within the gray matter was used for the SNR comparison. Therefore, in healthy volunteers, for both four-vessel and six-vessel scenarios, only the territories of the ICAs and VAs were included in the paired t-test, as the ECAs did not contribute to intracranial regions. The non-selective pairs were subtracted to generate a conventional perfusion image, which was utilized to create a gray matter mask by applying an empirical threshold factor of 0.3 times the 99th percentile intensity^12^. The SNR of each vascular territory was first calculated independently and then averaged. The VTI images were 4-dimensional data, with the fourth dimension being the vascular territory dimension for each targeting artery. Voxels exhibiting the largest signal across the fourth dimension and within the gray matter mask were identified as the dominant region for each territory, and the mean signal was calculated within these voxels. The mean SNR across all territories was calculated as the ratio of the mean signal to the standard deviation (SD) within a background region, with weighting applied based on the number of voxels within each territory. All methodologies were consistent with those reported in previous studies^12,13^ and more details were provided in **Figure S2**.

Similarly, the SNR was calculated for the VTI of Moyamoya patients and compared between the 5-and 8-average datasets.

#### 3.6.2 Relative Labeling Efficiency Maps

To demonstrate the efficiency of encoding at 7T after B_0_ inhomogeneity correction during each VEASL cycle, the relative labeling efficiency (*rLabEff*) maps were calculated for the Experiment 4 by normalizing the vessel-selective signal to the non-selective perfusion images as follows:

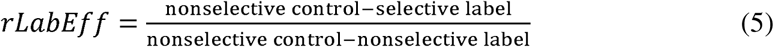

This indicated that *rLabEff* would equal 0 or 1 if the brain region were fully occupied by perfect control (non-inverted) or label (inverted) blood, respectively.

## 4 RESULTS

### 4.1 Theoretical SNR Efficiency Comparison

Compared to OES / IOES, MOGEN showed significant improvement in the theoretical SNR efficiency with reduced variability. Specifically, the SNR efficiency was improved from 0.827±0.038 to 0.844±0.014 (p < 0.001) for Para1 4Ves8Cycles, from 0.853±0.073 to 0.914±0.010 (p < 0.001) for Para2 4Ves8Cycles, from 0.642±0.084 to 0.721±0.071 (p < 0.001) for Para1 6Ves9Cycles, from 0.604±0.103 to 0.824±0.063 (p < 0.001) for Para2 6Ves9Cycles, from 0.671±0.074 to 0.721±0.064 (p < 0.001) for Para1 6Ves13Cycles and from 0.671±0.085 to 0.812±0.058 (p < 0.001) for Para2 6Ves13Cycles. Additionally, the MOGEN method demonstrated greater robustness to motion. As motion perturbations increased, the SNR efficiency of MOGEN remained higher than that of OES/IOES, as illustrated in Figure 3.

**Figure 3.**
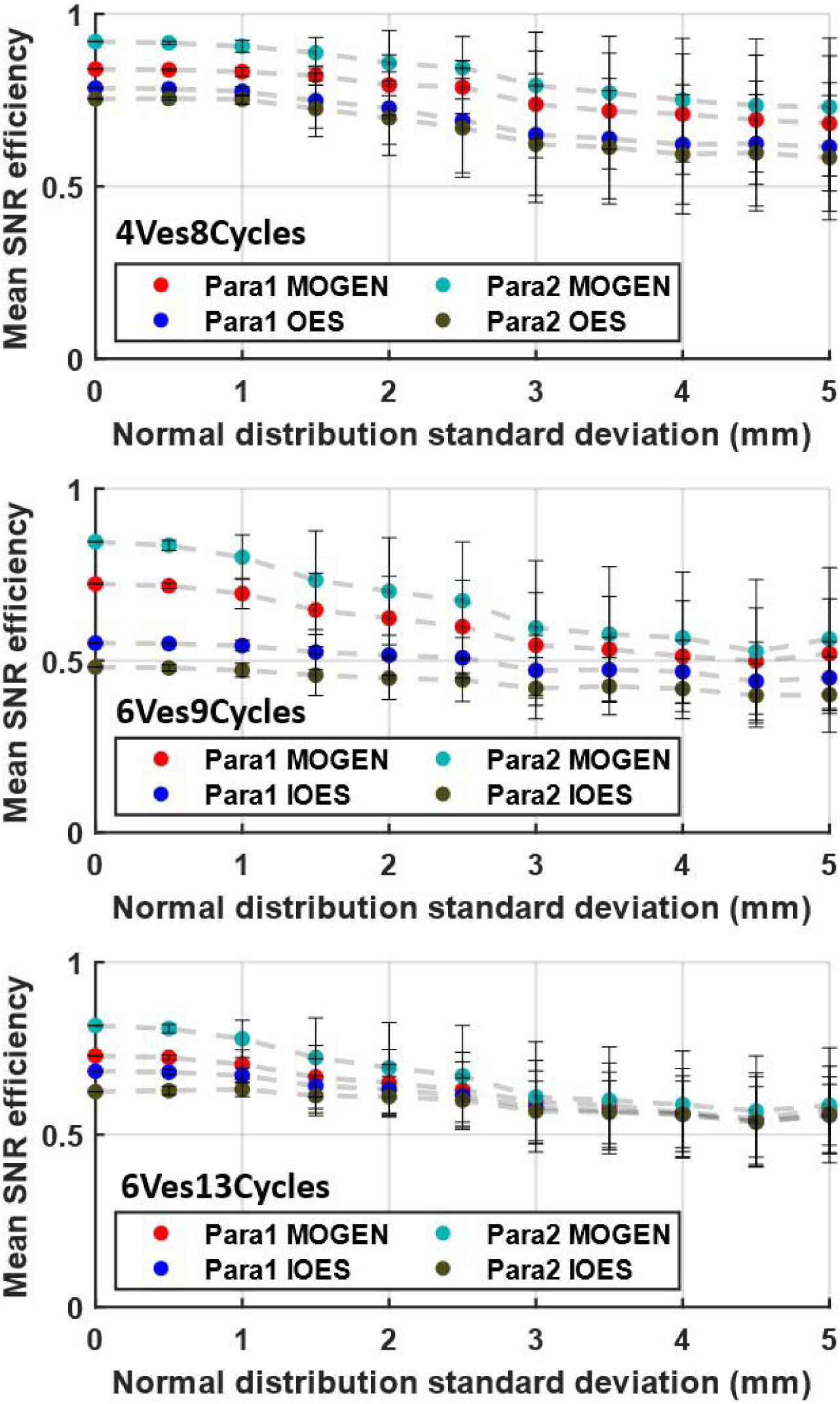
The mean SNR efficiency of MOGEN and OES/IOES in the simulation with motion. For either Para1 or Para2, MOGEN yielded a higher SNR efficiency than OES/IOES. In 6-artery encoding, the SNR efficiency of MOGEN is less dependent on the number of cycles than IOES.

### 4.2 In Vivo Mean SNR Comparison

In Experiment 1, one subject was excluded from the 6Ves13Cycles analysis due to significant head motion. Vessel-decoding failed in one subject each for 4Ves8Cycles and 6Ves13Cycles using IOES with Para2. In Experiment 2, three additional subjects were included for SNR comparison in the 6Ves13Cycles with Para2. As shown in **Figure S3**, using Para2, the SNR of MOGEN was significantly higher than OES by 15.4% for 4Ves8Cycles (p = 0.031) and higher than IOES by 19.1% for 6Ves13Cycles (p = 0.003). Using Para1, MOGEN showed a non-significant SNR improvement of 6.91% for 4Ves8Cycles compared to OES (p = 0.105) and 1.4% for 6Ves13Cycles compared to IOES (p = 0.777). MOGEN demonstrated more stable vessel-decoding, showing robustness even with six vessels in Figures 4a**, c, e**. The enhanced SNR in 2D MRA results allowed for better visualization of distal vessels in Figures 4b**, d, f**. In Experiment 2, the IOES performed poorly in 6Ves13Cycles for the second subject, completely failing in 6Ves9Cycles, as shown in **Figure S4b**.

**Figure 4.**
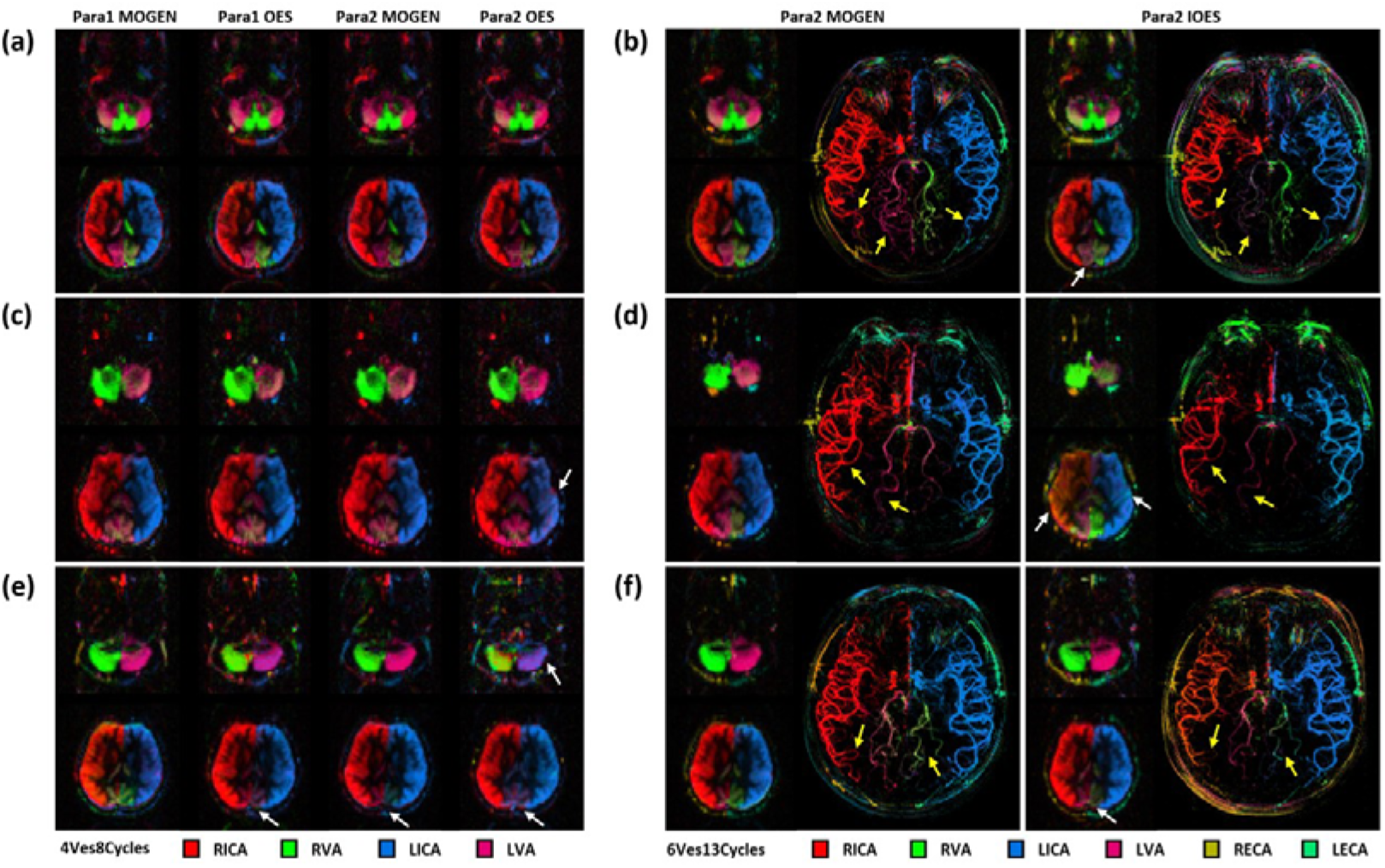
Three representative subjects from Experiment 1. (a, b) MOGEN exhibited better decoding in 6Ves13Cycles than IOES, with a clearer LVA supply area (white arrow) and more distinct distal vessels in MRA (yellow arrow), while no significant differences were observed for the 4Ves8Cycles scans. (c, d) The Para2 OES 4Ves8Cycles decoding was suboptimal (white arrow), and MOGEN performed significantly better in 6Ves13Cycles. (e, f) The RVA was decoded normally with high SNR in Para1 MOGEN 4Ves8Cycles, and distal vessels were also better visualized in 6Ves13Cycles. The clearer visualization of distal vessels indicated the SNR enhancement achieved with MOGEN.

For Moyamoya patients, the SNR was decreased by 21.8% when the number of averages was decreased from 8 to 5, but the average spatial correlation remained high at 0.93±0.028. Even for arteries with minimal SNR contribution to intracranial tissue, such as the non-dominant VAs and ECAs not yet establishing collateral pathway, the spatial correlation all remained above 0.72. Figure 5 showed that the visualization of collateral pathways had no significant change with only 5 averages, compared to that with 8 averages.

**Figure 5.**
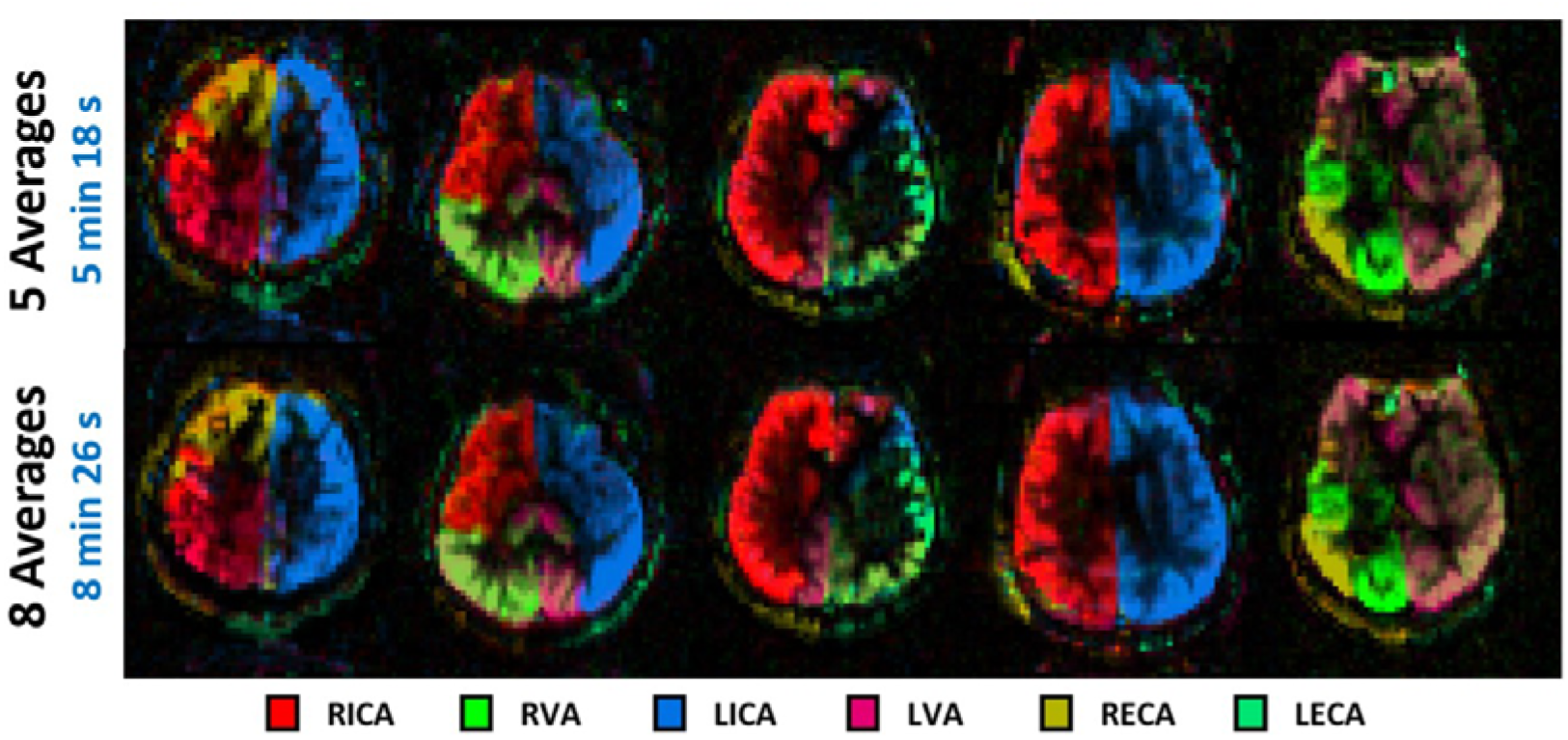
Typical VTI maps obtained with 5 averages and 8 averages using MOGEN from 5 MMD patients. When only 5 averages (with a total scan time of 5 min 18 s) were used, the decoding performance was generally similar to that with 8 averages (8 min 26 s), without a substantial compromise of the visualization of collateral pathways.

### 4.3 Capability for Achieving Single-Artery Selectivity in Angiography

The subtraction images obtained in Experiment 3 demonstrated that MOGEN_V2, incorporating 2D vessel size information, effectively displayed only the downstream branches of the artery of interest, minimizing interference from other arteries. The representative subject shown in Figure 6 supports the assertion that MOGEN_V2 not only achieved better single-artery selectivity, but also provided stronger signal in the distal branches of the posterior cerebral arteries (PCAs), which suggested that MOGEN_V2 could better avoid partial labeling and might overcome the typical suboptimal labeling issues (narrower labeling conditions using Para3) associated with the unipolar approach^19^. The vessel-encoded MRA data of other volunteers were shown in the supporting information **Figure S5**.

**Figure 6.**
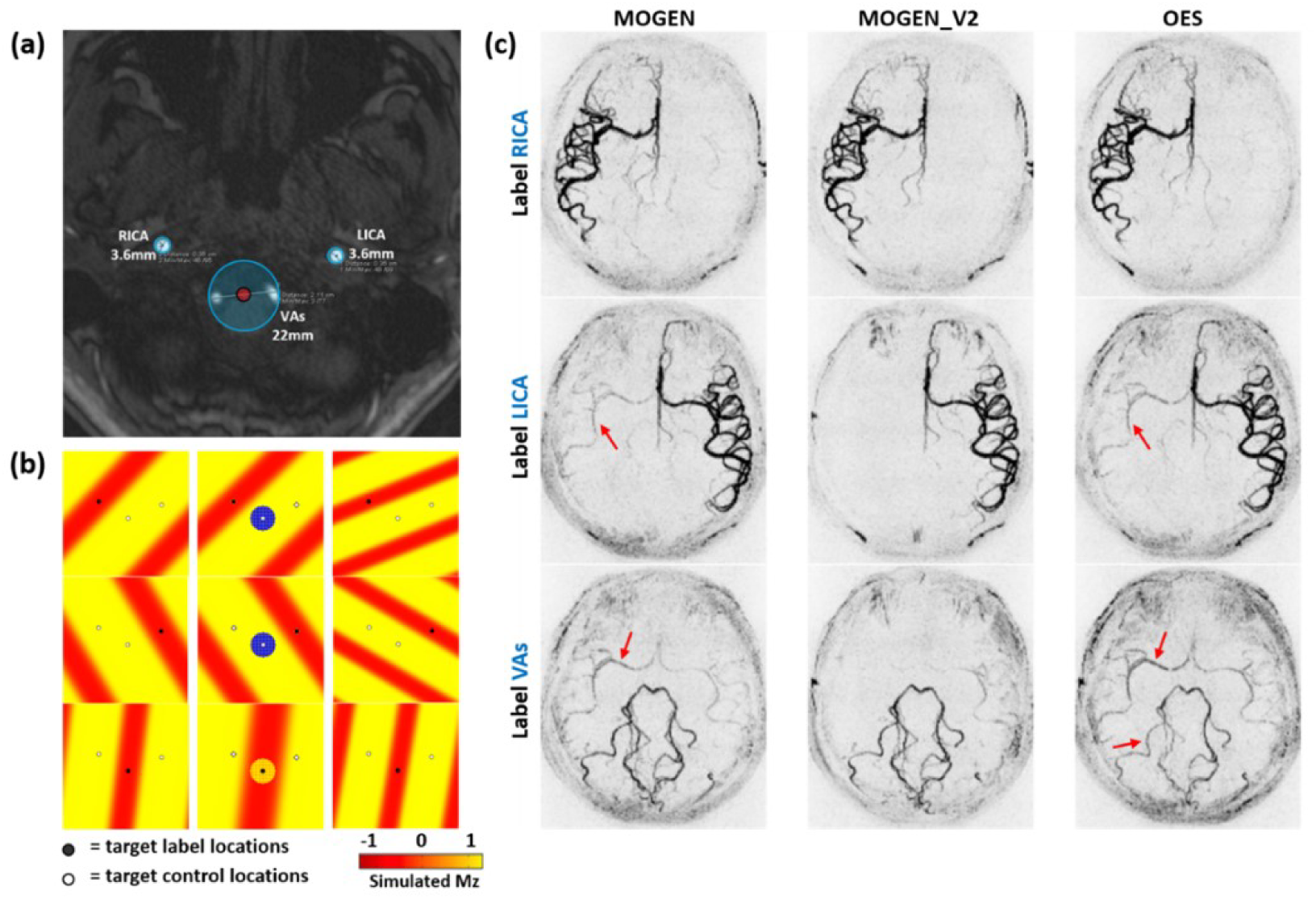
The encoding designs with MOGEN_V2 and the representative vessel-encoded MRA images from one healthy volunteer. (a) The vessel size was taken into account during the MOGEN calculation, with the vessels represented with circles. The two VAs was treated as a single labeling target and covered by a large circle. (b) The yellow dots around the vessel center indicate the label state, while the blue dots represent the control state. (c) MOGEN_V2 effectively avoids partial labeling associated with unipolar approaches in each cycle, with improved visibility of the distal arteries of RPCA compared to OES (red arrow).

### 4.4 The Off-Resonance Correction

In **Figure S6b**, the labeling regions (dark bands) were shifted away due to the presence of the off-resonance, but was precisely corrected and realigned to the desired encoding patterns using the MOGEN-based off-resonance correction. At 7T, the *rLabEff* maps were comparable between the dynamic shimming and the MOGEN method with off-resonance correction across seven subjects. Slight differences in *rLabEff* values and partial labeling effects were observed, as shown in Figure 7, but both methods effectively restored VEASL encoding and conveyed similar vascular territory information even without vessel-decoding. The results suggested that the MOGEN-based off-resonance correction might be suitable for applications at ultra-high fields, with the cost of only a 20 s field map.

**Figure 7.**
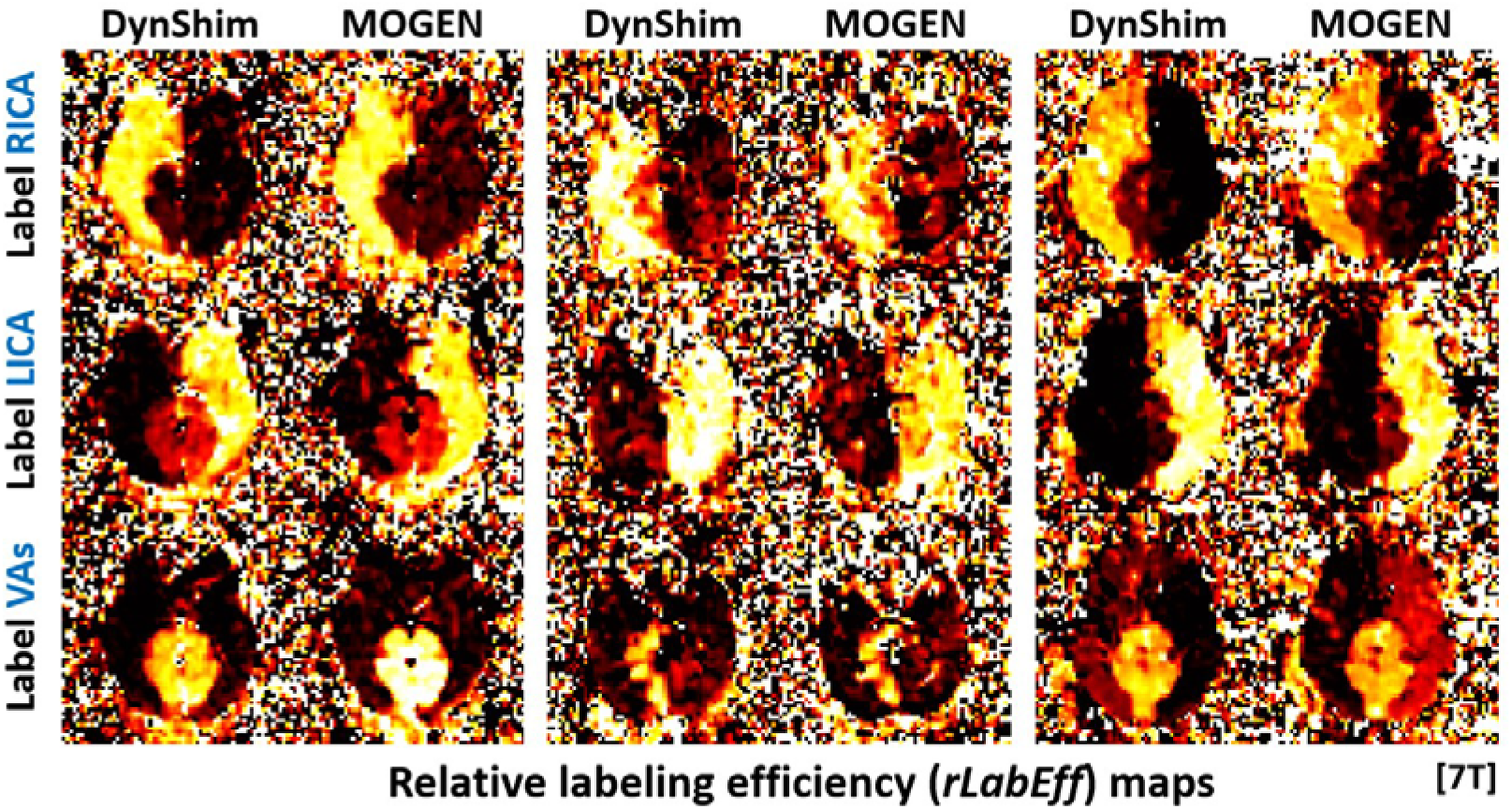
Representative *rLabEff* maps from three healthy volunteers obtained with dynamic shimming (DynShim) and MOGEN-based off-resonance correction at 7T. Although slight discrepancies in *rLabEff* values and partial labeling effects could be observed, both methods efficiently restored VEASL encoding.

## 5 DISCUSSION

In this study, the MOGEN method was proposed to account for the spatial modulation under arbitrary PCASL parameter settings to achieve optimal encoding for VEASL perfusion imaging and angiography. MOGEN has several benefits, including improving the flexibility of PCASL parameters selection for VEASL, high SNR efficiency, being applicable to any number of vessels and arrangements, and allowing consideration of vessel size. These benefits were demonstrated with simulation, phantom experiments, healthy volunteers experiments and patient experiments.

The key advantage of MOGEN lies in release of the assumption of cosine modulation in OES, and thus the constraints on the PCASL parameters. This enables dedicated optimization of the VEASL protocol for each specific application scenario. For instance, in cerebrovascular diseases, it is desirable to select PCASL parameters that are less sensitive to the variations of blood flow velocity between arteries or subjects. However, such PCASL parameters may result in a spatial modulation deviating from the cosine function. In the scenario where partial labeling is undesirable^19^, the unipolar approach with optimized parameter can be employed. However, this would also lead to a significantly non-cosine modulation^19^ and thus could benefit from MOGEN.

Vessel decoding failure occurred in one healthy volunteer using Para2 with OES/IOES in Experiment 1, reflecting potential instability in vessel-decoding for the two approaches when the spatial modulation substantially deviates from the cosine function, vessel positions change markedly, and the number of target vessels increases. Although IOES accounts for spatial modulation information during encoding, the decoding failure may still arise when such deviations become notable^14^.

The theoretical SNR efficiency of six-vessel encoding is generally lower than that of four-vessel encoding, as the ECAs and ICAs are often closely spaced above the carotid bifurcation. This region also exhibits significant variations in the arrangement of the four intracranial arteries, ICAs and VAs. The MOGEN method has been shown to better handle scenarios with more randomly distributed vessels compared to IOES, allowing for robust simultaneous encoding of intra-and extracranial arteries. We choose to place the labeling plane at slightly above the carotid bifurcation, instead of at the level of the V3 segment of the VAs that is commonly used in previous studies^13,22^. This is because the ECAs divide into multiple branches quickly after originating from the carotid bifurcation, and we want to label ECAs before the branching so as to better characterize the ECA perfusion in clinical application, e.g. assessment of collateral circulation from ECAs in Moyamoya diseases. Our patient data support that the MOGEN method using Para2 is a quite SNR-efficient strategy in Moyamoya disease, even at the carotid bifurcation, as vessel-decoding remains robust despite reduced the SNR of VEASL.

As for six-vessel encoding, both simulation (Figure 3) and in vivo results (**Figure S4**) indicated the performance of IOES was strongly dependent on the number of cycles, while the dependence for MOGEN was less distinct. In simulations using Para2, the mean SNR efficiency of MOGEN decreased from 0.824 to 0.812 when comparing 6Ves9Cycles to 6Ves13Cycles. Interestingly, as shown in **Figure S4a**, one subject even exhibited improved visualization of distal PCAs using MOGEN with 9 cycles, compared to that with 13 cycles. This implied that MOGEN might enhance the efficiency of selecting the optimal encoding combinations and was capable of yielding optimal SNR efficiency with minimum encoding cycles, which was beneficial for reducing the scan time. For six-vessel encoding, the scan time is less than 4 minutes using 5 averages and 9 cycles.

Figure 6 showed that incorporating the 2D vessel size information to MOGEN (i.e. MOGEN_V2) was useful for avoiding partial and suboptimal labeling and enhancing single-artery selective capabilities. For preliminary demonstration of the concept, each vessel was simply represented as a circle in the labeling plane in the experiment. In the subject shown in Figure 6a, we treated the two VAs as a single target. MOGEN_V2 can be treated as creating a circular protection zone around each vessel, meaning that precise vessel contours are not required. A slightly larger area than the vessel itself is sufficient. This concept may be particularly useful for distinguishing between the ECAs and ICAs. We plan to further validate this approach in future work.

This study successfully demonstrated the feasibility of the MOGEN-based off-resonance correction at 7T. At ultra-high fields, B_0_ inhomogeneities could lead to severe loss of perfusion signals^26^. The MOGEN-based off-resonance correction achieves *rLabEff* maps similar to dynamic shimming, although the principle of the two approaches is different. Dynamic shimming applies a first-order field correction over the labeling plane for all encoding cycles^23^. In contrast, MOGEN compensates for the off-resonance by changing the transverse gradient blips and RF pulse phases for each individual encoding cycle. Given the relatively smooth nature of the phase difference maps with minimal noise^21^, the measurement should be sufficiently robust. A limitation of MOGEN-based off-resonance correction is that it does not address through-plane off-resonance effects, whereas dynamic shimming can compensate for these^23^.

One major limitation of MOGEN is that the spatial modulation is dependent on the blood velocity, and the actual modulation may be different from the one used in the MOGEN calculation. However, this limitation also applies to other methods. OES necessitates that spatial modulation closely approximates a cosine function, a condition that is satisfied when using Para1 at a velocity around 30 cm/s^18^. Similarly, random encoding utilizes spatial modulation under a specific PCASL parameters setting to build a signal dictionary^17^. The advantage of MOGEN is that it offers greater flexibility in using the PCASL parameters that has a low sensitivity to velocity variations (e.g., Para2).

Head motion during VEASL scans may degrade encoding accuracy. We improved MOGEN’s robustness against motion from two aspects. First, the same cost function as in IOES was adopted in MOGEN, in which the minimum wavelength from the encoding matrix was penalized. Second, an exponential kernel with a specific decay rate was used to perform a weighted averaging of the dictionary of spatial modulation maps along the offset dimension. This kernel essentially can be considered a probability density function of the motion. Since the control-label transition or a narrow control (or label) band in the spatial modulation is more prone to be affected the smoothing processing, the algorithm will be guided to avoid placing the vessels at the edges of the transition and use a larger wavelength. However, it should be noted that in this study we did not extensively test the effect of this intuitive smoothing approach, and the MOGEN algorithm is expected to still work well without it.

## 6 CONCLUSION

This study proposed a novel method, i.e. MOGEN, to optimize the encoding design for VEASL by considering the spatial modulation. The simulations and experiments show that MOGEN has brought multiple important benefits for VEASL, including high SNR-efficiency, flexibility of PCASL parameters, easy consideration of the vessel size, and straightforward off-resonance correction. MOGEN is expected to largely improve the robustness and usability of VEASL in various applications.

## Data Availability

All data produced in the present study are available upon reasonable request to the authors.

## Notes

**Acknowledgments:** This work was supported by Natural Science Foundation of Shanghai (22ZR1403900) and National Natural Science Foundation of China (82302156). TO is supported by a Sir Henry Dale Fellowship jointly funded by the Wellcome Trust and the Royal Society (220204/Z/20/Z). For the purpose of open access, the author has applied a CC BY public copyright license to any Author Accepted Manuscript version arising from this submission.

### Competing Interest Statement

Thomas Okell is a co-author of a US patent relating to the VEASL Bayesian analysis method used in this work.

### Funding Statement

This work was supported by Natural Science Foundation of Shanghai (22ZR1403900) and National Natural Science Foundation of China (82302156). TO is supported by a Sir Henry Dale Fellowship jointly funded by the Wellcome Trust and the Royal Society (220204/Z/20/Z). For the purpose of open access, the author has applied a CC BY public copyright license to any Author Accepted Manuscript version arising from this submission.

### Author Declarations

The scans were performed under a technical development protocol agreed by the ethical committee of Fudan University. The patient study was approved by the ethical committee of the Second People's Hospital of Changzhou, the Third Affiliated Hospital of Nanjing Medical University. Written informed consentwas obtained from each subject, before participating in this study.

