## Supporting Information for "MOdulation-Guided ENcoding (MOGEN) Scheme for Vessel-Encoded Arterial Spin Labeling"

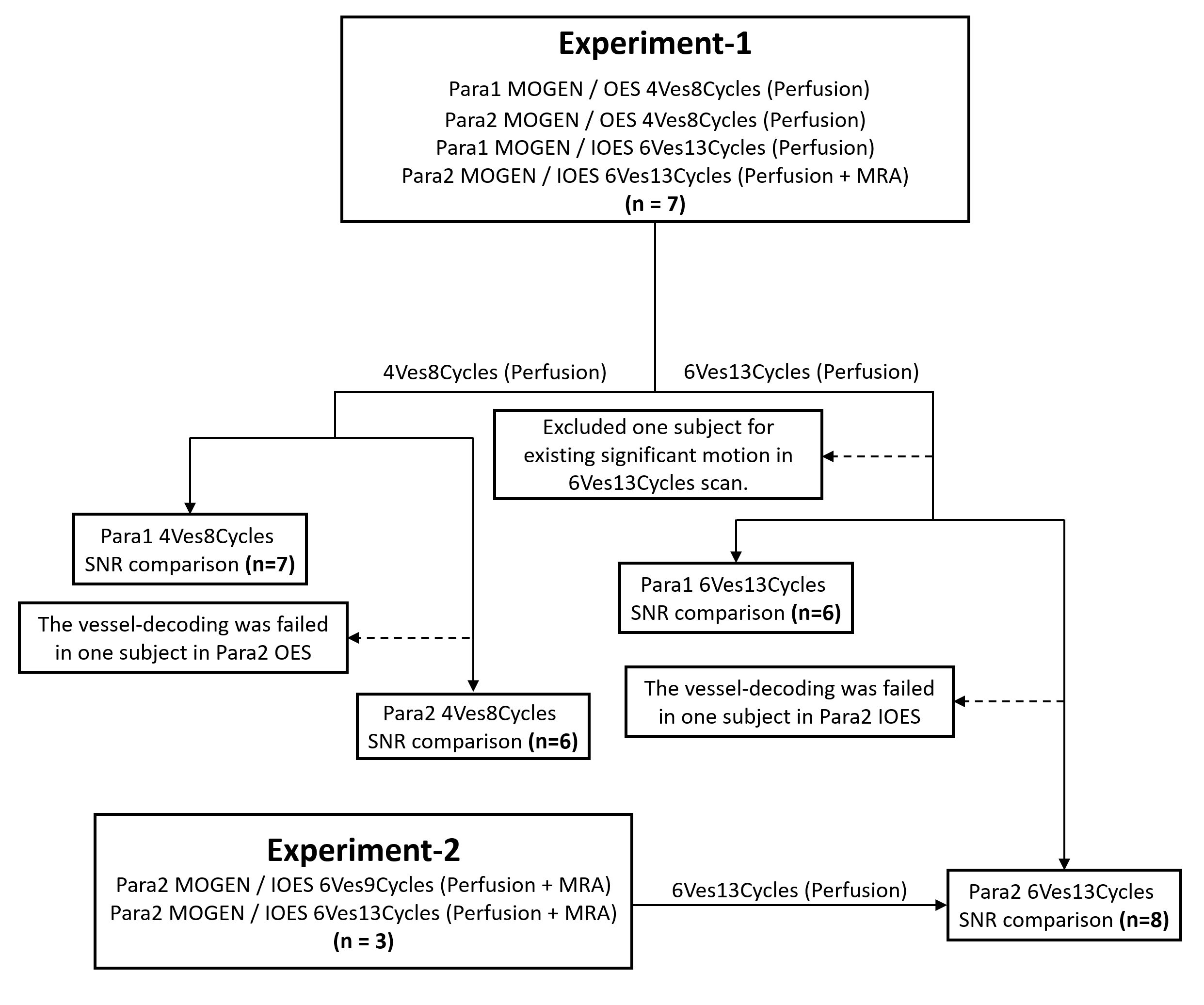

**Figure S1**. Flowchart detailing the subject inclusion process in the two experiments. Experiment 1: seven subjects were included, Para1 and Para2 were applied in 4Ves8Cycles and 6Ves13Cycles to compare the SNR between MOGEN and OES / IOES. One subject showed failed decoding in both OES and IOES with Para2, and thus was excluded from the statistical analysis. Experiment 2: three subjects were included for the six-vessel scenario with 6Ves9Cycles and 6Ves13Cycles using Para2. An additional scan of 2D dynamic vessel-encoded MRA was performed for the case with Para2 and six vessels. Statistical comparisons in SNR were performed only for the perfusion scans.

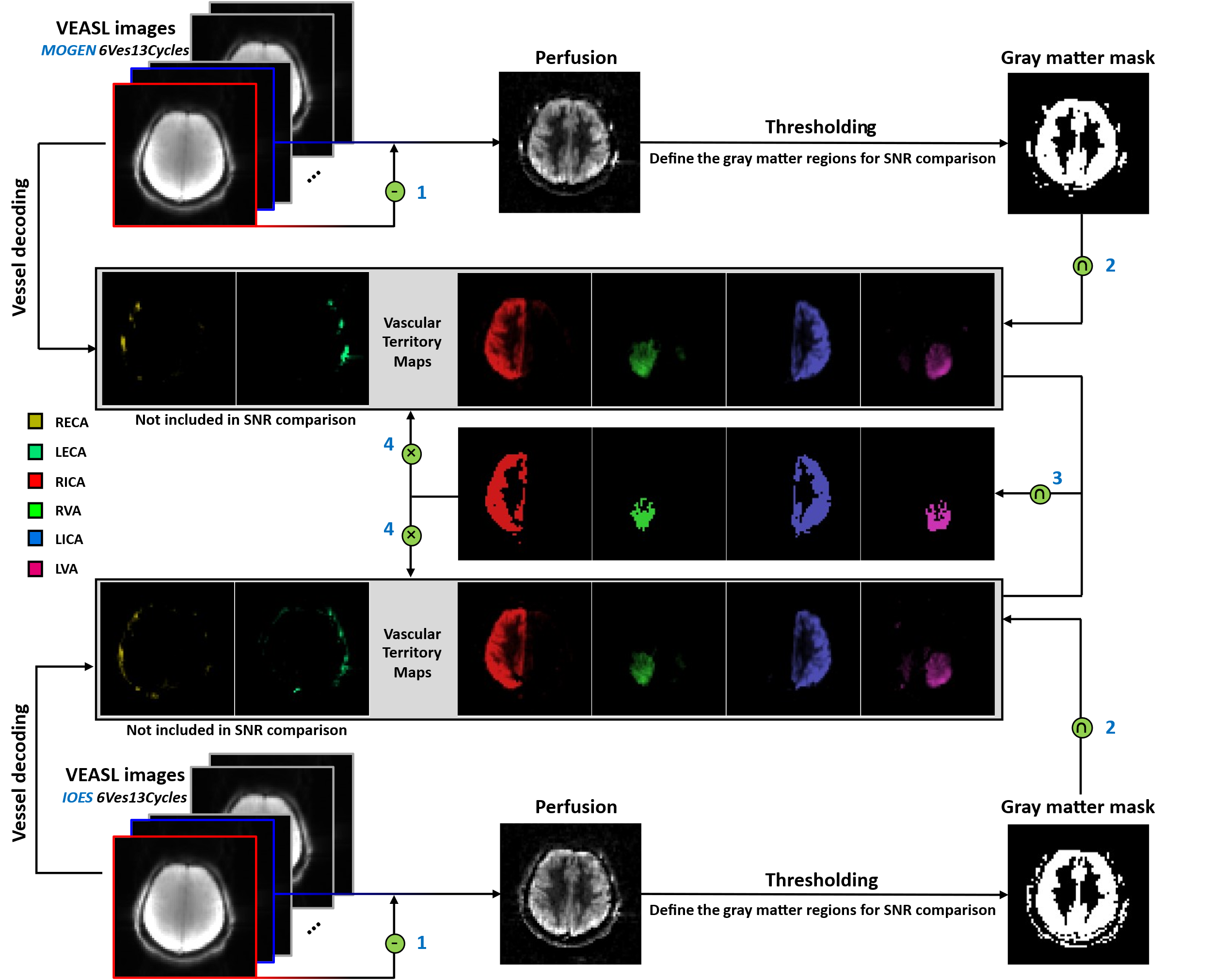

**Figure S2**. Procedure for calculating SNR of the in vivo VEASL perfusion data, exemplified with a 6Ves13Cycles scan. (1) The non-selective pairs were subtracted to generate perfusion maps and an empirical threshold were used to define the gray matter region; (2, 3) The decoded VTI images from MOGEN and IOES for each territory of ICAs and VAs were intersected with the gray matter to define the voxels with dominant supply for each artery; (4) The mean signal for each territory was extracted from the VTI images, weighted by the number of voxels in each territory, divided by the standard deviation of the background region, and then averaged to calculate the mean SNR.

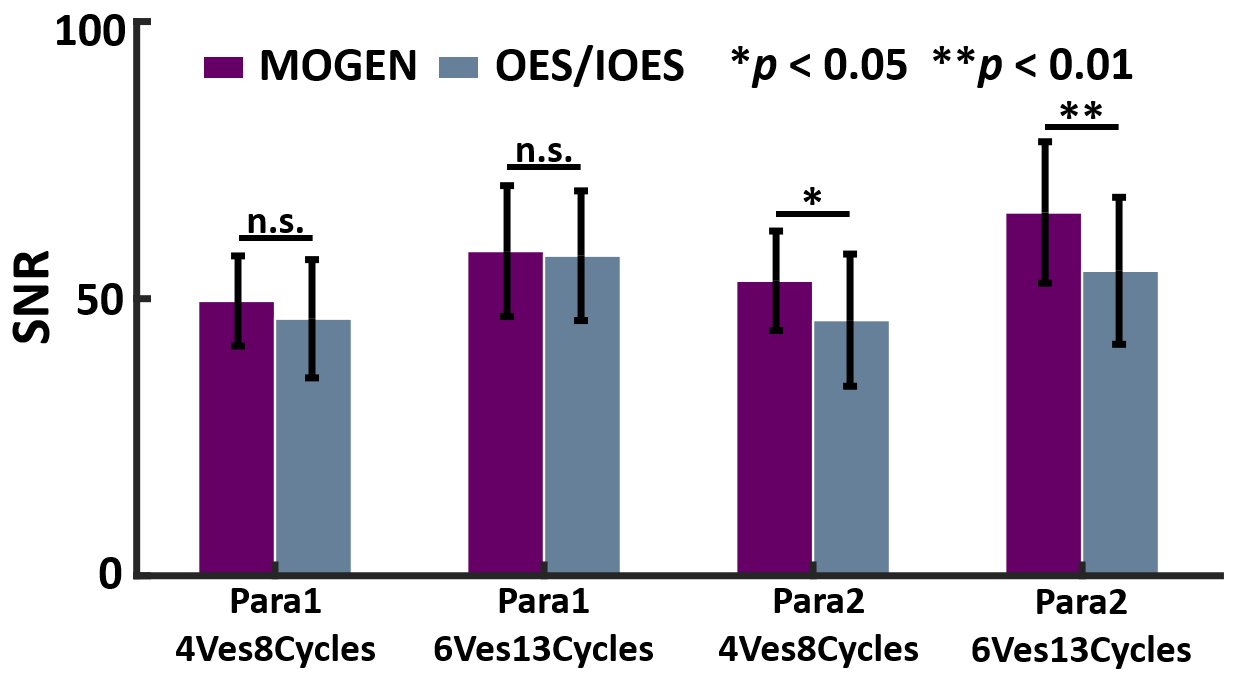

**Figure S3**. Comparison of mean SNR on healthy volunteers between MOGEN and OES/IOES under different PCASL parameters, number of vessels and number of encoding cycles. Using Para1, the SNR of MOGEN was higher than OES/IOES but not significant. Using Para2, the MOGEN method exhibited significantly higher SNR than OES/IOES.

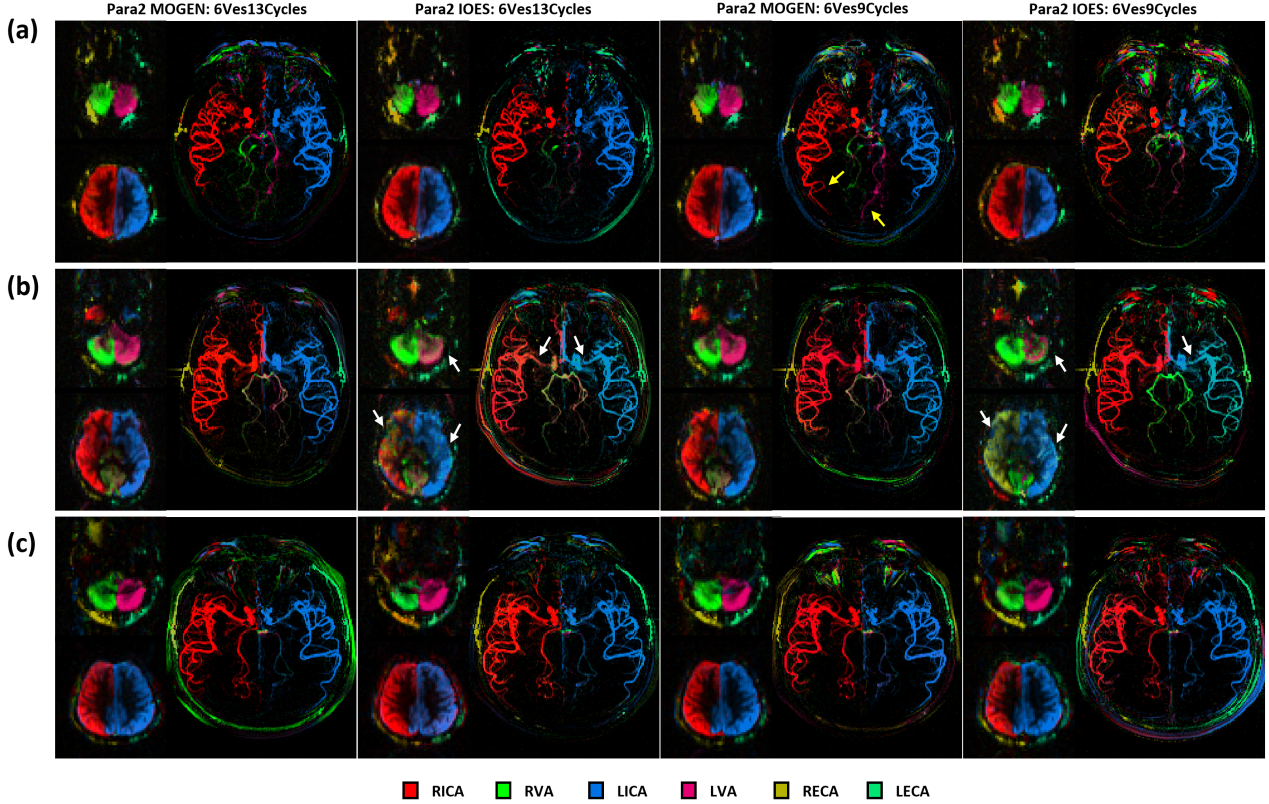

**Figure S4**. Comparison of the VTI maps between 13 and 9 cycles on three subjects from Experiment 2. For MOGEN, reducing cycles from 13 to 9 has little impact on vessel encoding. However, for IOES, as seen in the second row, more encoding cycles do contribute to improved encoding performance (white arrow). In the first row, MRA shows improved distal visualization in MOGEN with 6Ves9Cycles (yellow arrow), indicating that increasing the number of cycles does not necessarily enhance SNR efficiency in MOGEN.

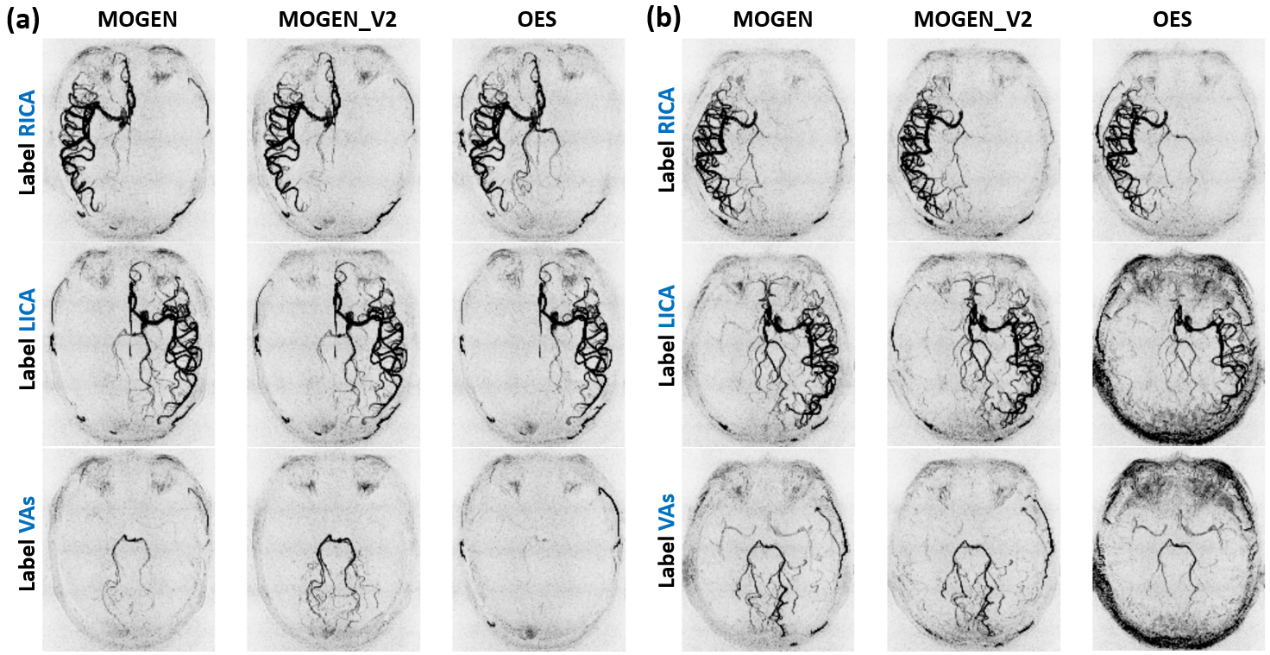

**Figure S5** Representative vessel encoded MRA images from two healthy volunteers.

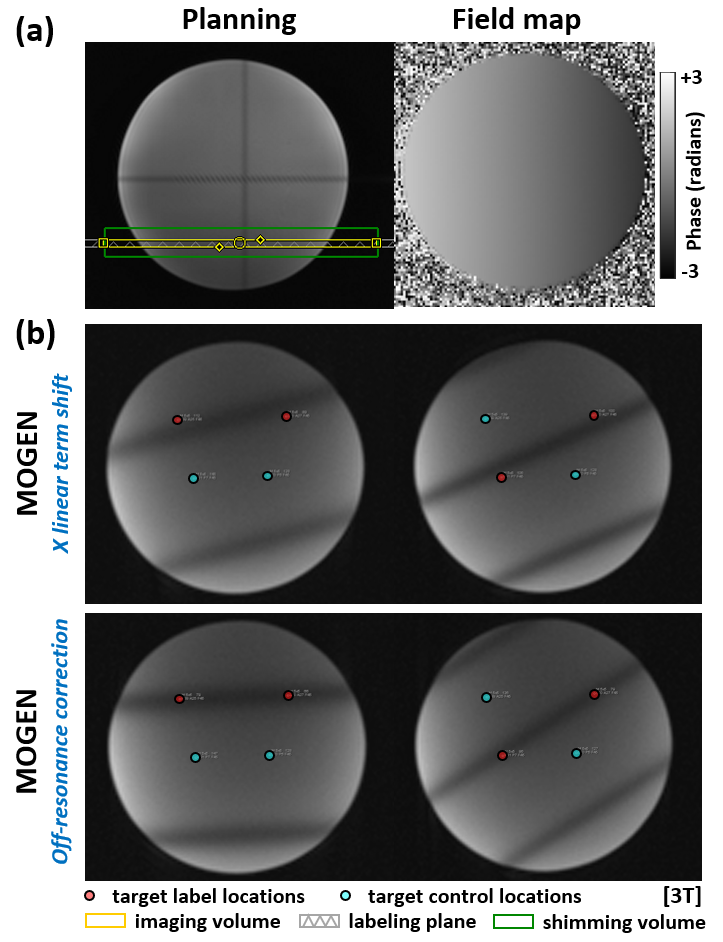

**Figure S6**. Illustration of the MOGEN-based off-resonance correction on phantom. (a) shows the locations of the labeling and imaging planes, as well as the measured field map. (b) shows two different encoding patterns before and after off-resonance correction. The encoding patterns were shifted due to off-resonance in the X direction and were corrected using the MOGEN-based off-resonance correction.
